# A New Paper Framework to Increase Reproducibility: Example Relating to Web Pharmacovigilance During COVID-19 in Italy

**DOI:** 10.1101/2022.05.03.22274607

**Authors:** Alessandro Rovetta, Lucia Castaldo

## Abstract

Reproducibility and transparency represent some of the main problems of scientific publishing. Currently, the editorial requests of academic journals and peer reviewers can divert the authors’ attention from an accurate description of the methods adopted, thus compromising these fundamental scientific aspects. This paves the way for the voluntary falsification of data to obtain striking results. Furthermore, the excessive expansion of introduction and discussion sections increases the likelihood of introducing evaluation bias. Since peer reviewers are generally unpaid for their work, they are not required to reproduce the analysis of the studies they review but only to assess methodological accuracy, reproducibility, and plausibility. Therefore, this paper aims to emphasize that the methods and results sections are the central parts of quantitative analysis. In this regard, we firmly believe that the peer review process should, whenever possible, reproduce the analysis from scratch. Consequently, authors must be required to provide a simple and straightforward tutorial to reproduce the analysis as it was conceived both methodologically and chronologically. Ideas, insights, and discussions among the authors must also be reported. This complete description can be provided as integrative material published with the main manuscript, which is nothing more than a summary of methods and results. Such a procedure would represent the first step to improving the quality of scientific publications, waiting for unscientific concepts such as “publish or perish” to be eradicated from the academic world. In this manuscript, we provide a framework that can serve as a fully reproducible and transparent example of analysis. The aim is to investigate the Italian netizens’ web interest in paracetamol, ibuprofen, and nimesulide from 2015 to 2022, searching for causal associations with the fever symptom and COVID-19. The infodemiological tool “Google Trends” has been used to collect the data. Correlational analysis showed plausible causal associations between paracetamol, ibuprofen, and fever due to seasonal flu and COVID-19 and, although to a minor extent, COVID-19 vaccines side effects. Paracetamol was the most historically searched substance. However, the trend of ibuprofen has caught up with that of paracetamol in 2022. Interest in paracetamol, ibuprofen, and nimesulide increased substantially during the COVID-19 pandemic period. We conclude that web pharmacovigilance via Google Trends can provide relevant evidence for monitoring drug intake in relation to epidemiologically significant events such as epidemics and mass vaccination campaigns.

## Introduction

The introduction of a scientific paper is designed to introduce the reader to the topic covered in the manuscript. In particular, this section should contain the minimum amount of information essential to contextualize the paper in the current scenario. In this regard it is necessary to consider that, unless it is purposefully informative, a study is intended for an audience of expert researchers who do not need to be introduced to the subject except if there is very recent evidence to discuss. Furthermore, it is unrealistic to believe that some introductory pages can provide a thorough background to a public that is not already well versed in the matter. Therefore, the choice to deepen this section should be left entirely to the paper’s authors according to their necessities. On the contrary, a brief description of the present scientific situation and study aim should always be furnished. At best, a list of preliminary concepts – necessary for a complete understanding of the outcomes – could be reported to facilitate the reader; in this case, the latter is invited to consult the suggested references. By doing so, we firmly believe that authors can focus more on a detailed description of a manuscript’s milestones, that is, its methods and results. Indeed, regarding these last two sections, a peer reviewer is currently not required to check the findings quantitatively but only to evaluate their plausibility and the theoretical correctness of the approach used (e.g., model’s assumptions) [1, 2]; obviously, this drastically decreases the peer review effectiveness and the publications’ quality [1, 3]. At the same time, judgments on more discursive sections such as introduction and discussion are more likely to be influenced by personal biases [1]. To overcome these problems, the authors of this paper suggest two complementary solutions: i) pay peer reviewers for their work (as it should be for any other worker) so that they can ensure more thorough scrutiny, and ii) realize manuscripts focusing primarily on the methods’ reproducibility and less on scientifically secondary issues such as a broad introduction or discussion, so as to facilitate the replication of the results. In this way, authors can also save time and resources by concentrating their efforts merely on the quality of the evidence presented.

According to the above, in this paper we investigate the web interest in drugs such as paracetamol, ibuprofen, and nimesulide (PIN) from 2015 to 2022 in Italy. These substances are typically used to counteract symptoms such as fever, myalgia, headache, and other inflammations accompanying respiratory tract infections and otorhinolaryngological diseases [4-6]. Since such manifestations are typical of common cold, seasonal flu, and COVID-19, the aim was to monitor users’ historical interest in PIN in order to assess possible causal relationships with seasonal flu, COVID-19 trends, and other epidemiologically relevant events (e.g., vaccinations) as well as differences in their use and/or knowledge over the years. On this point, it is crucial to consider that paracetamol and ibuprofen are over-the-counter drugs, while nimesulide must be prescribed by the attending physician.

### Preliminary concepts

COVID-19 pandemic in Italy [7-9], infodemiology and infoveillance [10], seasonal flu in Italy [11, 12].

## Methods

This section is typically meant to describe in detail the methodological approach of the study so as to ensure full reproducibility and transparency. However, given the tight editorial demands regarding the maximum number of figures, tables, and words, authors often struggle to make the methods sufficiently clear and complete. For instance, it is not uncommon for editors to require authors to shorten sentences and leave out auxiliary specifications to scale text within editorial standards. By doing so, the risk of compromising or unnecessarily complicating reproducibility increases significantly. While the supplementary files can serve as a potential solution to the problem, in some fields or subfields, it is still rare to find all the necessary specifications to carry out the analysis from scratch [13, 14]. This lack opens the way not only to bona fide errors but also to voluntary falsifications. For this reason, the authors of this paper suggest the following design: the “Methods” section should contain a descriptive summary of the methods adopted in order to allow the reader a complete understanding of the results. A detailed description, including the data collection process, choice of tests, verification of models’ assumptions, and similar aspects, should be provided as integrative material. By doing so, the reading of the paper would be smooth and clear, and, at the same time, it would be possible to increase the results’ reproducibility by adding details that could be omitted or explained too briefly due to editorial stylistic requirements. In particular, the concept of integrative material is philosophically opposed to that of supplementary material: indeed, this is not an additional material anymore but an essential tool for the validity of the manuscript. It must be understood that the integrative material should form the core of any scientific work and that the manuscript is built around it. The simplest way to ideally represent this section is that of a tutorial for other researchers to explain thoroughly and faithfully each step of the analysis carried out by the paper’s authors. Furthermore, we would like to underline an often underestimated issue: as highlighted by Greenland et al., the authors’ biases are part the entire data analysis procedure, starting from the data collection and/or the choice of test hypotheses [15]. For this reason, we believe that the integrative material should chronologically describe not only the complete scientific procedure but also the data interpretation, ideas and research questions born after the paper’s conception and then added to the latter, rejected data, negative results (to be summarized in the main manuscript), and the like.

According to the above, in this paper we adopted Google Trends [16] to perform a retrospective longitudinal pharmacovigilance analysis in Italy. In particular, we collected the web relative search volume (RSV) of the topics “Fever,” Paracetamol,” “Ibuprofen,” and “Nimesulide” within specific periods from January 2015 to April 2022. We examined RSV trends and related queries before and after 2020 in order to discover and comprehend eventual causal associations with epidemiological phenomena such as seasonal flu and COVID-19. In this way, we were also able to check for any confounding factors. Based on the latter, additional analyses were subsequently introduced into the study (i.e., the search for causal relationships between PIN and COVID-19 vaccines). Since the datasets were generally non-normal and large (N>30), we looked for Spearman correlations and Welch ANOVA/t-test significant differences between all the keywords or between the RSVs of a single keyword during different timelapses (we exploited the central limit theorem). P-value was used as a graded measure of the dataset’s compatibility with the test hypothesis.

## Results

This section should present the analysis results on a quantitative level. It is essential to adopt a schematic and simple presentation. The discursive part should be limited to explaining the meaning of the findings concisely. Thus, whoever needs to use such data will be able to do it without complications whatsoever. Exact P-values should always be reported in the **Integrative section** to allow for meta-analyses or adjustments. Negative results must be signaled.

According to the above, we found marked and significant correlations between Fever (x), Paracetamol (y), and Ibuprofen (z) web topics from 2015 to 2022 (r_xy_=0.79, 95% CI [0.67, 0.86], P_xy_<.0001; r_xz_=0.61, 95% CI [0.45, 0.74], P_xz_<.0001; r_yz_=0.88, 95% CI [0.80, 0.93], P_yz_<.0001). On the contrary, the web topic Nimesulide seemed generally unrelated to the others except for some spikes (**Figure 1**).

**Figure 1.**
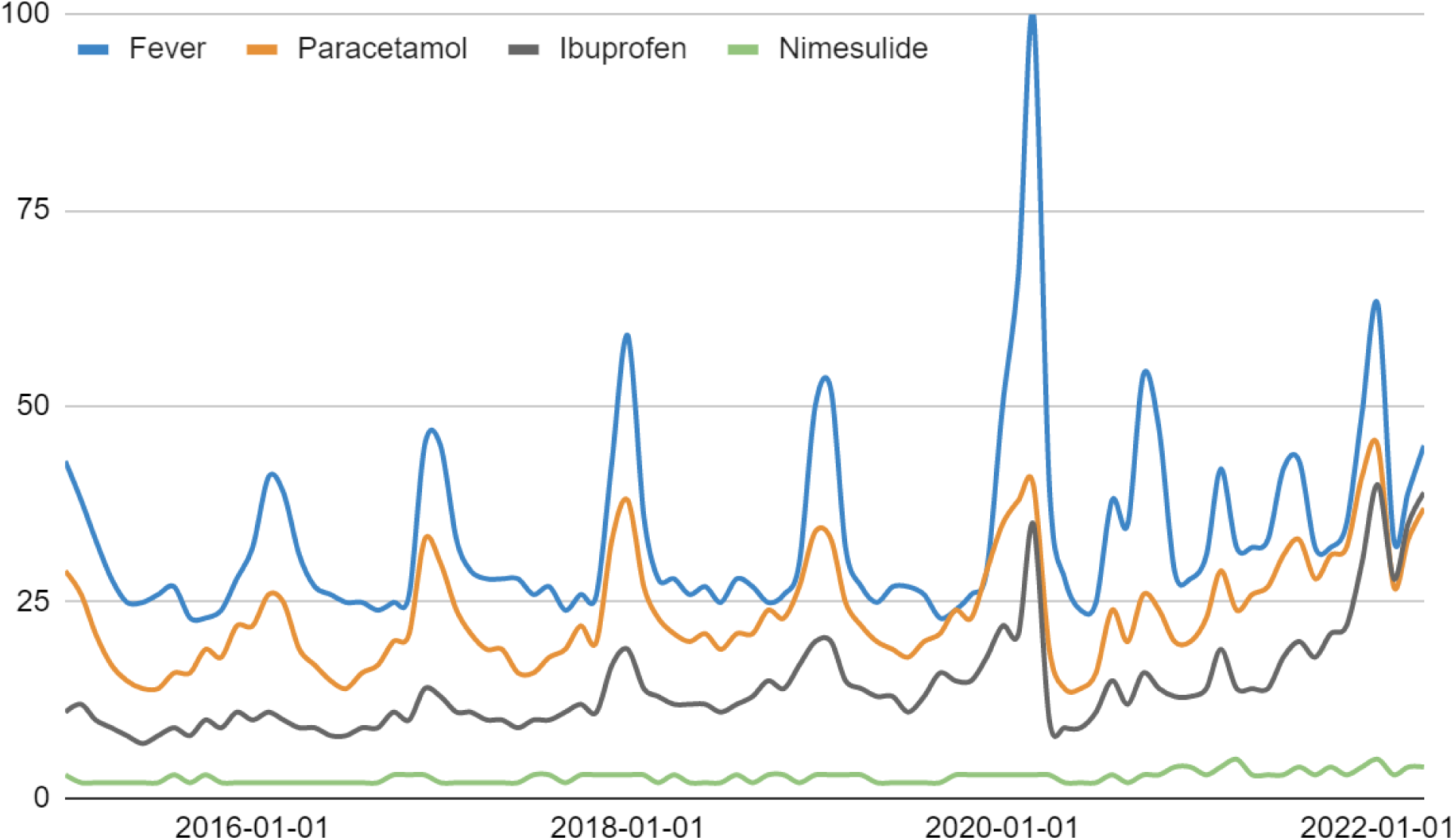
Fever, paracetamol, ibuprofen, and nimesulide monthly web interest in Italy from January 2015 to April 2022. Before 2020, the peaks always coincided with flu season (nimesulide excluded). During COVID-19, anomalies are evident.

During the pandemic period January 2020 - April 2022 (**Figure 2**, daily RSVs instead of monthly RSVs), web searches for ibuprofen matched those for paracetamol. Besides, out-of-season peaks (e.g., August and October-November 2020) and the absence of winter 2020-2021 peaks can be observed. The latter phenomenon can be explained by an unusually low incidence of seasonal flu. Considering the top and rising related queries, we deduced that the media hype influenced the searches on fever more than those on the two drugs (**Integrative section**). In particular, the total RSV of the keywords containing the terms “coronavirus” or “covid” was 58 for fever, 13 for paracetamol, and 10 for ibuprofen. Analyzing the rising queries, we found a majority of searches related to COVID-19 (83% for fever, 61% for paracetamol, and 56% for ibuprofen) and a minority related to COVID-19 vaccines (17% for fever, 33% for paracetamol, and 6% for ibuprofen). Again, we found marked and significant correlations between Fever (x1), Paracetamol (y1), and Ibuprofen (z1) (r_x1y1_=0.78, 95% CI [0.70, 0.85], P_x1y1_<.0001; r_x1z1_=0.62, 95% CI [0.47, 0.74], P_x1z1_<.0001; r_y1z1_=0.89, 95% CI [0.82, 0.93], P_y1z1_<.0001). Welch ANOVA and t-test analysis confirmed a substantial difference between the volumes of these queries (P<.0001).

**Figure 2.**
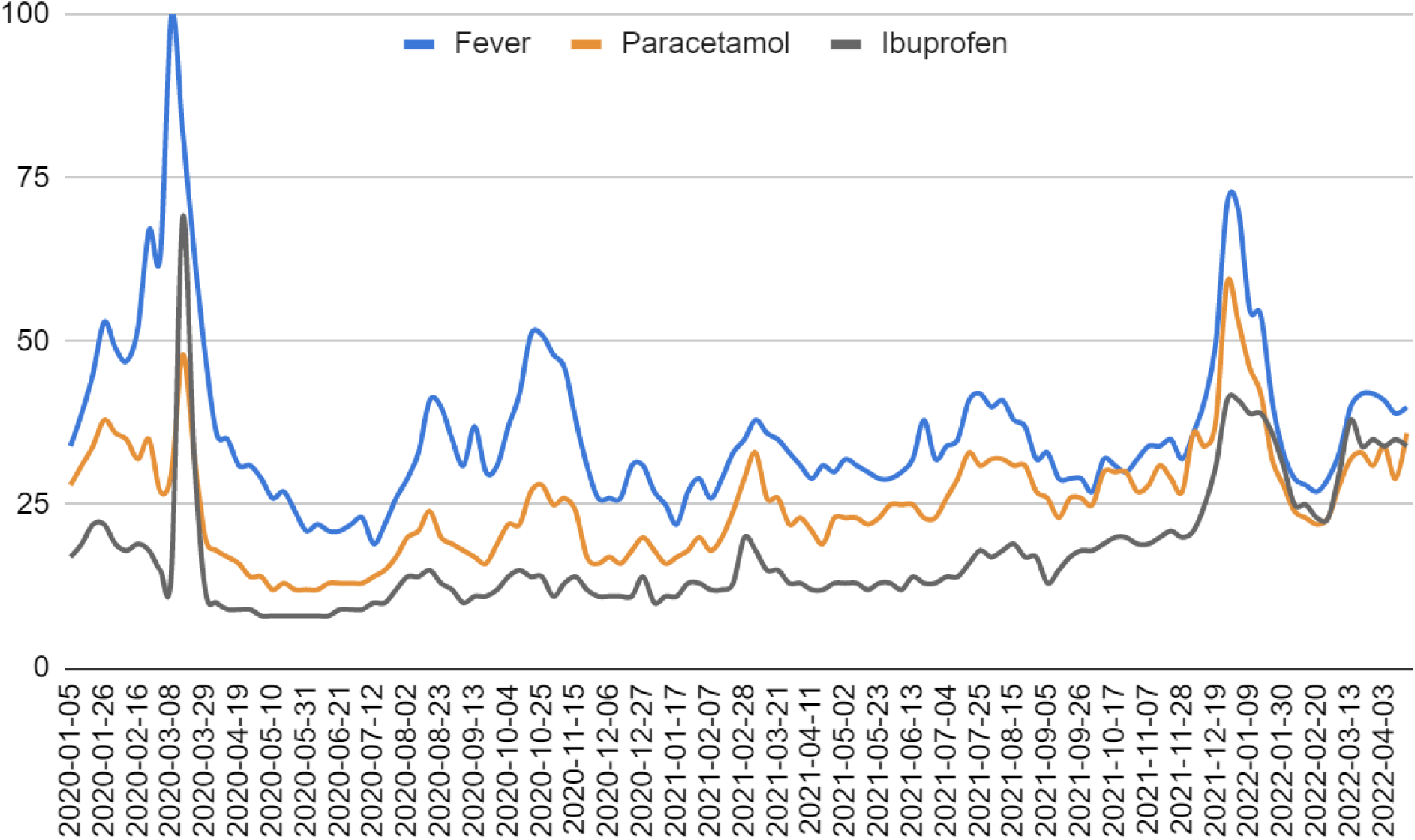
Fever, paracetamol, and ibuprofen weekly web interest in Italy from January 2020 to April 2022.

As shown in **Figure 3**, there has been a strong association between searches for these substances and COVID-19 vaccines. Indeed, the web search volumes are perfectly aligned with national vaccinations. Nimesulide-related RSV was negligible (**Integrative section**). By calculating the correlations between COVID-19 vaccinations (x2), “tachipirina vaccino” query (y2), and “brufen vaccino + ibuprofene vaccino” query (z2), we obtained marked and significant values (r_x2y2_=0.88, 95% CI: [0.78, 0.93], P_x2y2_<.0001 and r_x2z2_=0.85, 95% CI: [0.72, 0.91], P_x2z2_<.0001). The average RSV were 16 (SD 19) for Paracetamol (X2) and 3 (SD 4) for Ibuprofen (Y2). Welch t-test confirmed the significance of the difference (P<.0001).

**Figure 3.**
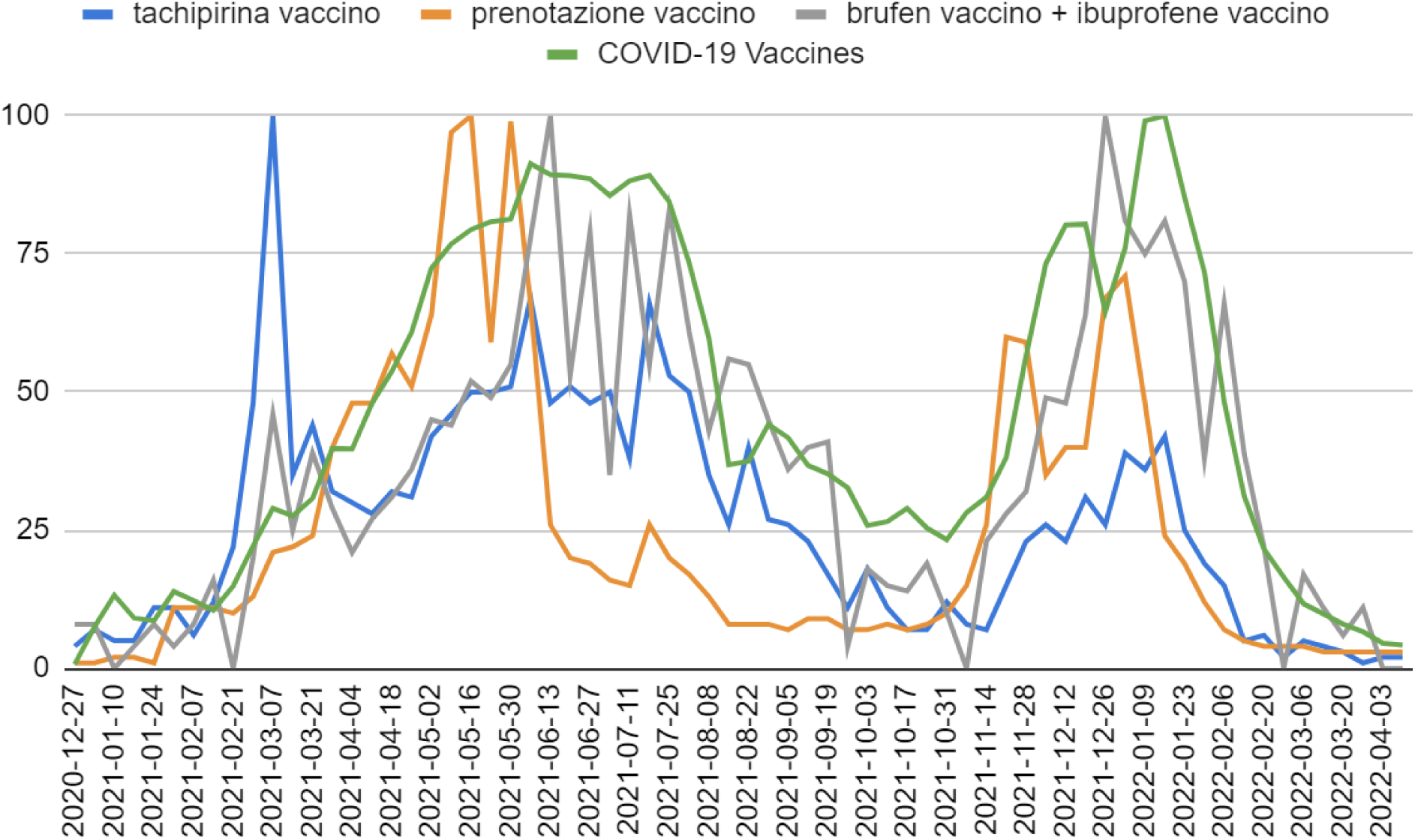
COVID-19 vaccinations and vaccine-related queries in Italy from January 2021 to April 2022. The volumes of each query have been downloaded separately and put in this graph (therefore, no quantitative information can be deduced from this graph). Nimesulide-related RSV was negligible.

The “Nimesulide” web topic registered a marked and significant increase from October 2020 onwards, preceded by a drop after the first COVID-19 wave (May – July 2020). Indeed, considering that the dataset showed no trend (**Figure 4**), the Welch t-test returned a significant difference (P<.0001). The average values were 47 (SD 5) before October 2020 and 69 (SD 13) after October 2020. Top and rising queries confirmed the direct association between these searches and COVID-19 during the pandemic period (**Integrative section**).

**Figure 4.**
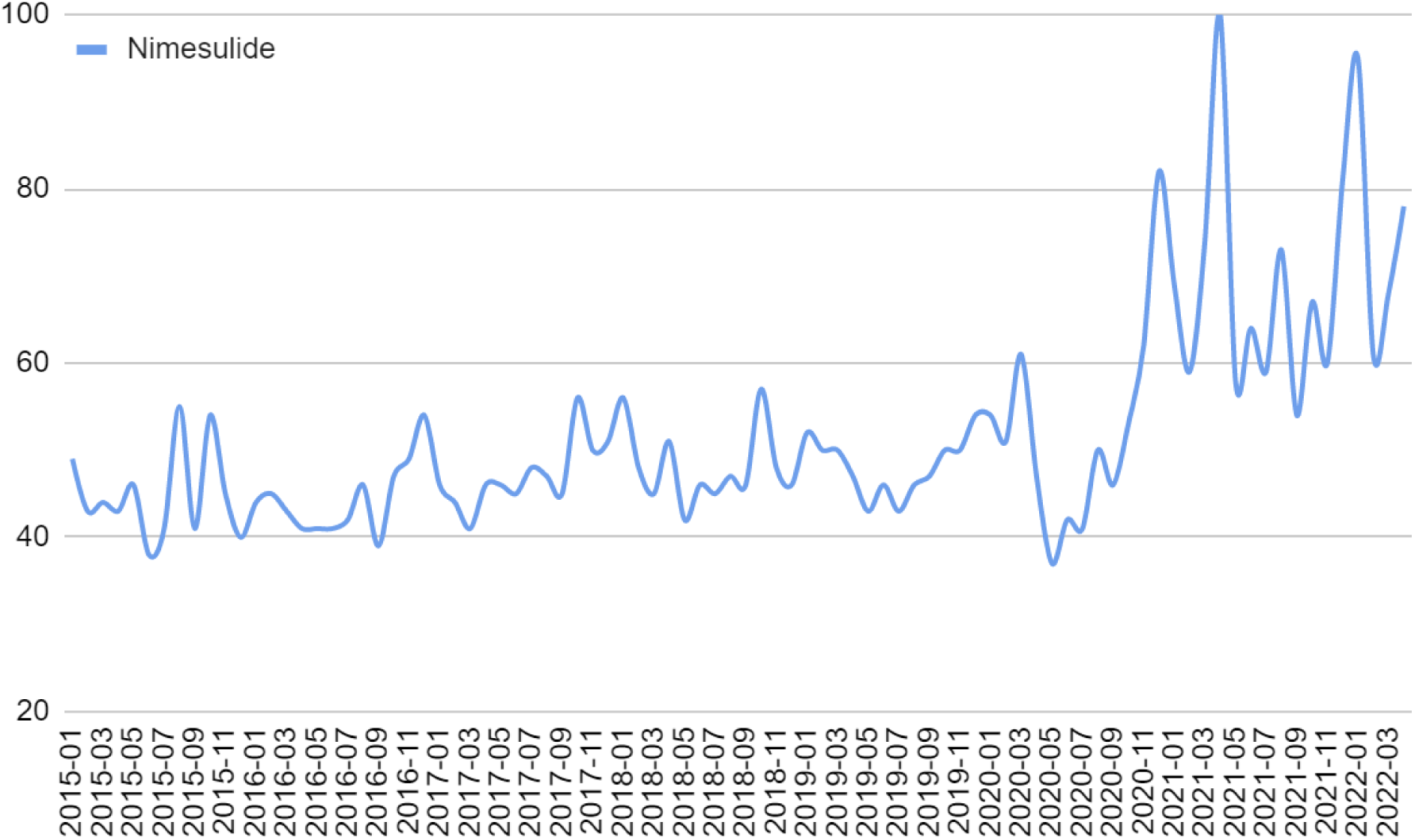
Nimesulide monthly web interest in Italy from January 2015 to April 2022.

## Discussion

This section often consists of three or four main sections, such as i) a summary of the paper’s main findings, ii) a comparison with other literature, iii) limitations (and strengths), and iv) conclusions. In this regard, the authors of this paper consider sections i), iii), and, eventually, iv) to be appropriate. In particular, comparison with other literature should be optional for the same reasons set out in the Introduction section plus one: unless systematic reviews or meta-analyses are performed, such comparisons are likely to be undermined by authors’ biases and are statistically unreliable since no combining techniques (e.g., Fisher formula for P-values) can be employed. Indeed, a scientist must always admit that biases are part of any analysis as they represent a cognitive feature of our human nature [17]. These can be limited or – more or less – managed by quantitative analyses but certainly not eliminated. Hence, it is extremely difficult to conduct a neutral comparison without exploiting the methods of systematic reviews and meta-analyses [18]. For these reasons, we suggest leaving any comparisons optional according to the authors’ aims rather than forcing them to disclose their biases when not necessary. Furthermore, we suggest presenting the results with caution, avoiding overstatements, and trying to describe in detail what was found rather than convince the reader of the study’s relevance. Finally, we believe that this is only a first step towards the qualitative improvement of scientific evidence. Specifically, academia too often pushes researchers into dishonesty in order to obtain relevant funding or positions [19]. Ergo, as a scientific community, we must rethink this system in order to prevent such inadmissible situations.

According to the above, in this paper we found marked and significant correlations between Fever, Paracetamol, and Ibuprofen web topics in Italy. Such associations have historically been evident both from 2015 to 2022 (monthly RSV) and during the COVID-19 pandemic (weekly RSV). Since paracetamol and ibuprofen are typically adopted to counteract common cold, seasonal flu, and COVID-19 symptoms such as fever, we have considered such relationships to be causal. Further evidence of causality is provided by Google Trends’ rising and top queries, which testify to a direct medical association between these terms. Besides, the web search peaks until 2020 coincided with the flu period. Ergo, Google Trends could be adopted by health authorities and research institutes to investigate drug interest (and intake) when linked to well-targeted hypotheses (e.g., the correlation between drug and symptoms typically treated with the drug). The search volumes increased substantially with the advent of COVID-19. Although there was evidence that such growth was mainly due to the disease, we have found plausible causal associations with confounding factors such as vaccines. In this regard, this study provides additional proof of the goodness of Google Trends in monitoring the vaccination status. Specifically, Google Trends could be adopted to scrutinize the online pharmacological interest linked to the adverse events of vaccines (or other drugs widely administrated). Indeed, the above scenario is plausibly causal since substances such as paracetamol and ibuprofen are recommended to treat the mild and moderate adverse reactions of COVID-19 vaccines [20, 21]. These findings also suggest that paracetamol could be the most used drug to counteract common COVID-19 vaccine-related side effects. Finally, we found a marked and significant increase in web interest in nimesulide during COVID-19. In particular, we detected a first anomalous peak during the first wave (spring 2020) and substantial growth in search volumes from the second wave (October 2020) onwards. Since Google Trends-related queries confirmed the direct association between these searches and COVID-19, and nimesulide is adopted to treat some typical COVID-19 symptoms, we consider these findings plausibly causal. No clear longitudinal correlation was found between nimesulide and the other keywords and topics examined.

## Limitations

Like all quantitative analyses, this paper is also limited in the choice of tests and hypotheses by the authors’ biases and eventual confounding factors. Statistical analysis cannot by itself provide definitive evidence of causality. Furthermore, we did not verify that the web interest was associated with a real intake of the drug examined. Finally, the Italian Google-related internet penetration limits the investigated public to about 70-75% of the population (age groups have not been considered).

## Data Availability

All data produced in the present work are contained in the manuscript

## Acknowledgments

I thank Dr. Luca Tramontana and Dr. Simone Raho for sharing and supporting the intent of this paper.

## Integrative section

We have collected the relative search volumes (RSV) as shown in **Table I1**. These keywords were selected to monitor the relationship between fever and paracetamol, ibuprofen, and nimesulide before and during COVID-19.

**Table I1.** Fever, Paracetamol, and Ibuprofen exact queries.

| Collection | Topic | Region | Period | Category | Search | URL |
| --- | --- | --- | --- | --- | --- | --- |
| C1 | Fever (Medical condition) | Italy | 1/1/15 - 4/22/22 | Health | Web Search | URL-1 |
|  | Paracetamol (Medication) |  |  |  |  |  |
|  | Ibuprofen (Medication) |  |  |  |  |  |
|  | Nimesulide (Drug) |  |  |  |  |  |
| C2 | Fever (Medical condition) | Italy | 1/1/20 - 4/22/22 | Health | Web Search | URL-2 |
|  | Paracetamol (Medication) |  |  |  |  |  |
|  | Ibuprofen (Medication) |  |  |  |  |  |
| C3 | Nimesulide (Drug) | Italy | 1/1/15 - 4/22/22 | Health | Web Search | URL-3 |

Regarding C1, considering that the “Nimesulide” RSV was low compared to others (**Figure I1**), we have defined the following vectors in RStudio software (version 4.2.0., https://www.rstudio.com/):

*x <-*

*c(43,38,33,28,25,25,26,27,23,23,24,28,32,41,39,31,27,26,25,25,24,25,26,45,45,33,29,28,28,28,26,27,24,26,26,43,59,36,28,28,26,27,25,28,27,25,26,30,50,52,32,27,25,27,27,26,23,24,26,29,50,67,100,40,28,24,25,38,35,54,47,29,28,31,42,32,32,33,42,43,32,32,35,49,63,33,39,45)* (Fever RSV, Collection 1),

*y <-*

*c(29,26,21,17,15,14,14,16,16,19,18,22,22,26,25,19,17,15,14,16,17,20,21,33,30,24,21,19,19,16,16,18,19,22,20,33,38,27,23,21,20,21,19,21,21,24,23,27,34,33,25,22,20,19,18,20,21,24,23,29,35,38,40,19,14,14,16,24,20,26,24,20,20,23,29,24,26,27,31,33,28,31,32,41,45,27,33,37)* (Paracetamol RSV, Collection 1)

*z <-*

*c(11,12,10,9,8,7,8,9,8,10,9,11,10,11,10,9,9,8,8,9,9,11,10,14,13,11,11,10,10,9,10,10,11,12,11,17,19,14,13,12,12,12,11,12,13,15,14,17,20,20,15,14,13,13,11,13,16,15,15,18,22,21,35,10,9,9,11,15,12,16,14,13,13,14,19,14,14,14,18,20,18,21,22,30,40,28,35,39)* (Ibuprofen RSV, Collection 1).

Using the Shapiro-Wilk test, we verified that the datasets were strongly non-normal (P_x_=2.596e-11, P_y_=.0001724, P_z_=1.425e-10). The following commands were used: *shapiro*.*test(x), shapiro*.*test(y)*, and *shapiro*.*test(z)*. Therefore, we calculated the Spearman correlation. We found marked and significant correlations between the topics (r_xy_=0.79, 95% CI [0.67, 0.86], P_xy_<2.2e-16; r_xz_=0.61, 95% CI [0.45, 0.74], P_xz_=3.331e-10; r_yz_=0.88, 95% CI [0.80, 0.93], P_yz_<2.2e-16). The following commands were used:

*cor.test(x,y,method=“spearman”,exact=FALSE), spearman.ci(x, y, nrep = 1000, conf.level = 0.95) cor.test(x,z,method=“spearman”,exact=FALSE), spearman.ci(x, z, nrep = 1000, conf.level = 0.95), cor.test(y,z,method=“spearman”,exact=FALSE), spearman.ci(y, z, nrep = 1000, conf.level = 0.95)*.

Since the drugs above are indicated to counteract seasonal flu or cold symptoms such as fever, we have considered these correlations to be causal.

**Figure I1.**
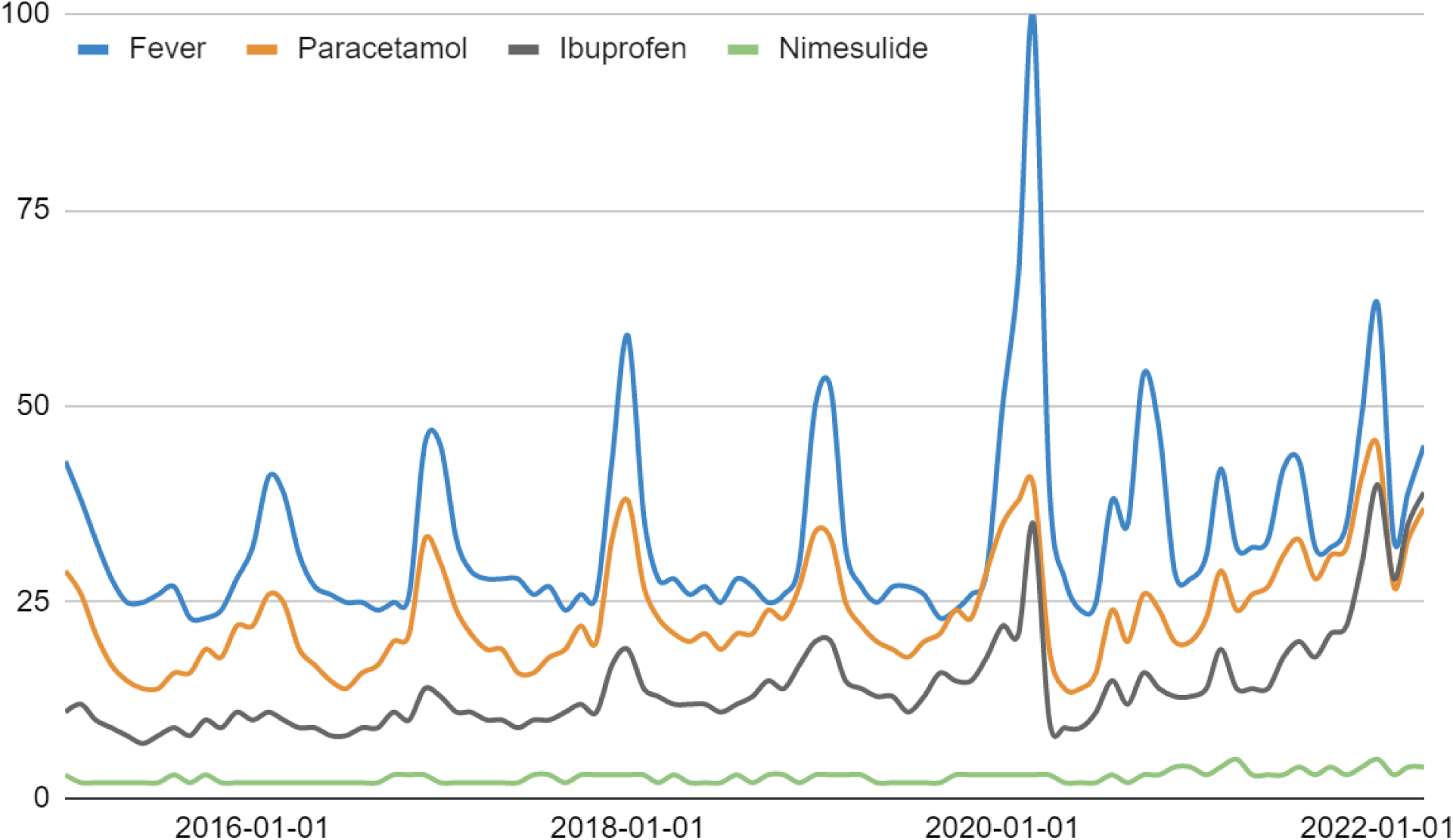
Fever, paracetamol, ibuprofen, and nimesulide monthly web interest in Italy from January 2015 to April 2022. Before 2020, the peaks always coincided with flu season (nimesulide excluded). During COVID-19, anomalies are evident.

Since the central limit theorem guarantees that the distribution of mean values is approximately normal for sufficiently large datasets (e.g., N>30), we employed a Welch one-way ANOVA to verify the significance of the differences between web interest in these topics (exact commands: *df <-data.frame(group = rep(c(‘A’,’B’, ‘C’), each=88), score = c(x,y,z))* and *oneway.test(score ∼ group, data = df, var.equal = FALSE)*). We obtained P<2.2e-16. The mean values were the following: mean RSV_x_ 34 (SD_x_ 12), mean RSV_y_ 24 (SD_y_ 7), mean RSV_z_ 14 (SD_z_ 7). One-tailed Welch t-test for single pairs returned very significant differences (P_xy_=2.252e-10, P_xz_<2.2e-16, P_yz_<2.2e-16). The following commands were used: *t.test(x, y, alternative = c(“greater”)), t.test(x, z, alternative = c(“greater”)), t.test(y, z, alternative = c(“greater”)), mean(x), sd(x), mean(y), sd(y), mean(z), sd(z)*.

As for C2, we have defined the following vectors:

*x1 <-*

*c(34,39,45,53,49,47,52,67,63,100,82,64,48,36,35,31,31,29,26,27,24,21,22,21,21,22,23,19,22,26,29,33,41,40,35,31,37,30,31,37,42,51,51,48,46,38,31,26,26,26,31,31,27,25,22,27,29,26,29,33,35,38,36,35,33,31,29,31,30,32,31,30,29,29,30,32,38,32,34,35,41,42,40,41,38,37,32,33,29,29,29,27,32,31,30,32,34,34,35,32,36,41,50,71,70,55,54,41,33,29,28,27,29,33,40,42,42,41,39,40)* (Fever RSV, Collection 2),

*y1 <-*

*c(28,31,34,38,36,35,32,35,27,30,48,33,20,18,17,16,14,14,12,13,12,12,12,13,13,13,13,14,15,17,20,21,24,20,19,18,17,16,19,22,22,27,28,25,26,24,17,16,17,16,18,20,18,16,17,18,20,18,20,24,29,33,26,26,22,23,21,19,23,23,23,22,23,25,25,25,23,23,26,29,33,31,32,32,31,31,27,26,23,26,26,25,30,30,30,27,28,31,29,27,36,34,38,59,53,46,42,32,28,24,23,22,23,28,32,33,31,34,29,36)* (Paracetamol RSV, Collection 2)

*z1 <-*

*c(17,19,22,22,19,18,19,18,15,15,69,33,12,10,9,9,9,8,8,8,8,8,8,8,9,9,9,10,10,12,14,14,15,13,12,10,11,11,12,14,15,14,14,11,13,14,12,11,11,11,11,14,10,11,11,13,13,12,12,13,20,18,15,15,13,13,12,12,13,13,13,12,13,13,12,14,13,13,14,14,16,18,17,18,19,17,17,13,15,17,18,18,19,20,20,19,19,20,21,20,21,25,31,41,41,39,39,36,31,25,25,23,23,30,38,34,35,34,35,34)* (Ibuprofen RSV, Collection 2).

Using the Shapiro-Wilk test, we verified that the datasets were strongly non-normal (P_x_=7.501e-11, P_y_=6.671e-05, P_z_=3.796e-12). The following commands were used: *shapiro.test(x1), shapiro.test(y1)*, and *shapiro.test(z1)*. Therefore, we calculated the Spearman correlation. We found marked and significant correlations between the topics (r_x1y1_=0.78, 95% CI [0.70, 0.85], P_x1y1_<2.2e-16; r_x1z1_=0.62, 95% CI [0.47, 0.74], P_x1z1_=6.058e-14; r_y1z1_=0.89, 95% CI [0.82, 0.93], P_y1z1_<2.2e-16). The following commands were used:

*cor.test(x1,y1,method=“spearman”,exact=FALSE), spearman.ci(x1, y1, nrep = 1000, conf.level = 0.95) cor.test(x1,z1,method=“spearman”,exact=FALSE), spearman.ci(x1, z1, nrep = 1000, conf.level = 0.95), cor.test(y1,z1,method=“spearman”,exact=FALSE), spearman.ci(y1, z1, nrep = 1000, conf.level = 0.95)*.

The web topics trends can be observed in **Figure I2**. It can be seen that searches for ibuprofen matched those for paracetamol during 2022. Besides, out-of-season peaks (e.g., August and October-November 2020) and the absence of winter 2020-2021 peaks can be observed. The latter phenomenon can be explained by an unusually low incidence of seasonal flu.

**Figure I2.**
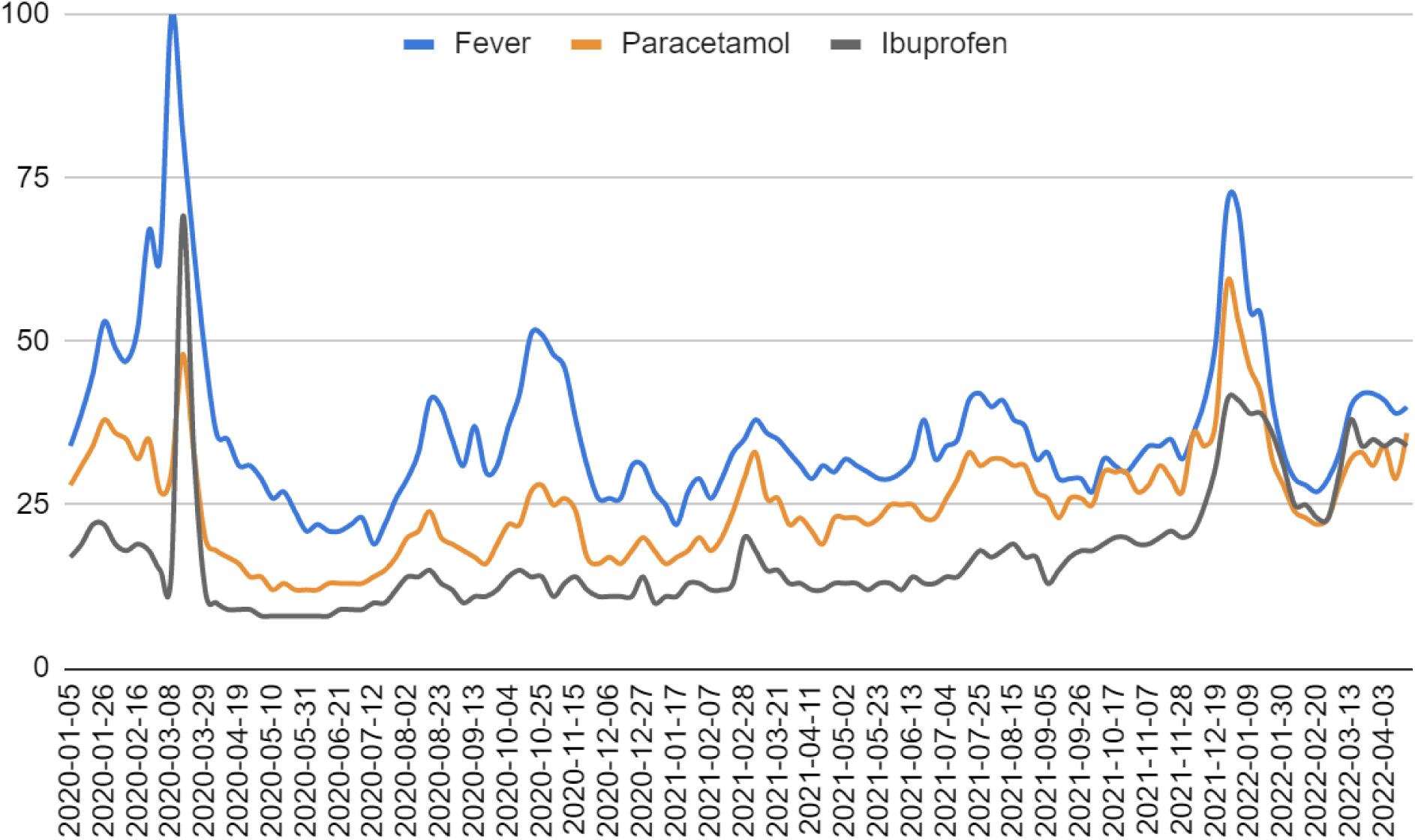
Fever, paracetamol, and ibuprofen weekly web interest in Italy from January 2020 to April 2022.

Observing the top and rising related queries, we deduced that the media hype influenced the searches on fever more than those on the two drugs (**Table I2**).

**Table I2.**
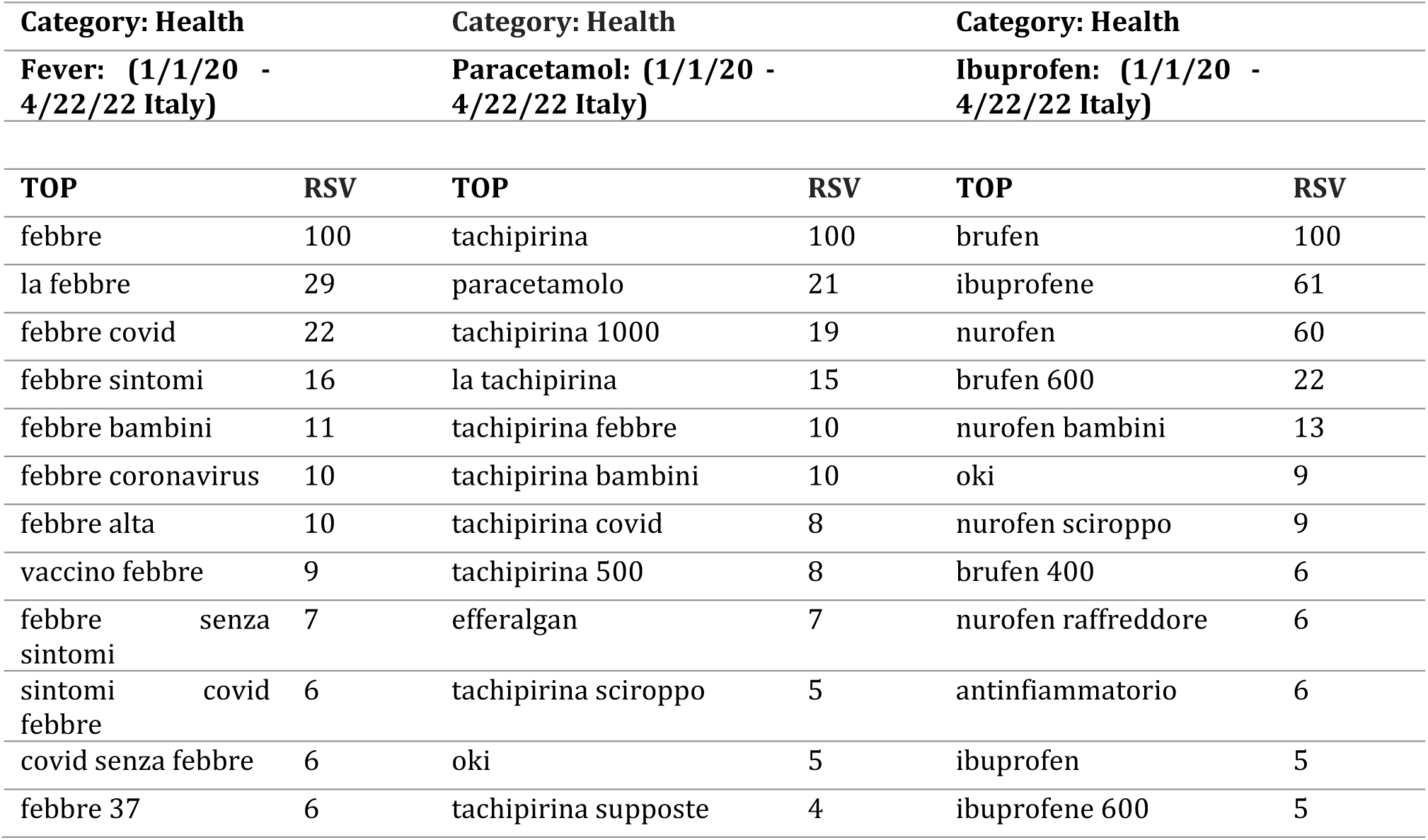
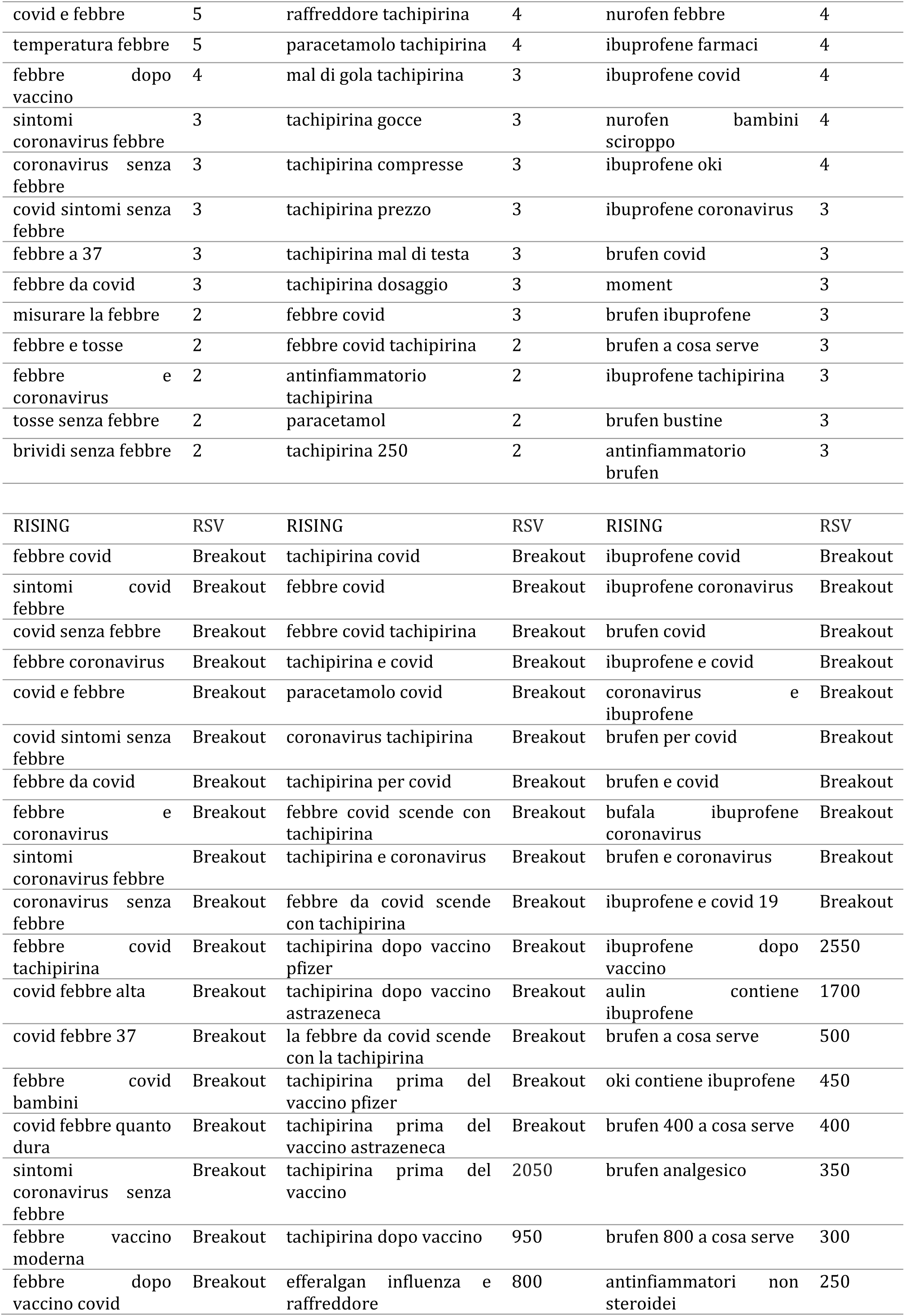
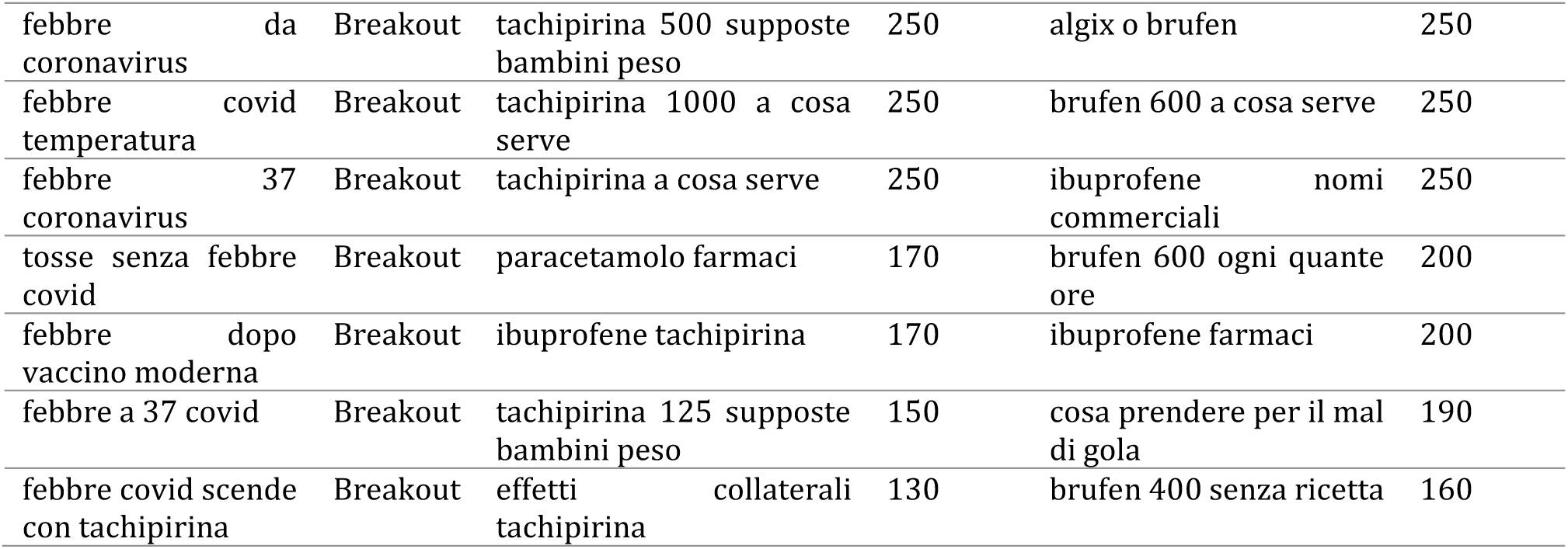
Fever, Paracetamol, and Ibuprofen top and rising queries.

In particular, the total RSV (sum) of keywords containing “coronavirus” or “covid” was 58 for fever, 13 for paracetamol, and 10 for ibuprofen. Analyzing the rising queries, we found a majority of searches related to COVID-19 (83% for fever, 61% for paracetamol, and 56% for ibuprofen) and a minority related to COVID-19 vaccines (17% for fever, 33% for paracetamol, and 6% for ibuprofen). This finding prompted us to think that vaccines could be an additional confounding factor (in addition to seasonal flu and colds) when searching for a causal association between COVID-19 incidence and paracetamol/ibuprofen. Therefore, we searched for the following keywords (**Table I3**) and compared them with normalized COVID-19 vaccinations (max value 100 and the other scale proportionally, https://github.com/owid/covid-19-data/blob/master/public/data/vaccinations/country_data/Italy.csv):

**Table I3.** COVID-19 vaccine-related exact queries.

| Collection | Exact keyword | Region | Period | Category | Search | URL |
| --- | --- | --- | --- | --- | --- | --- |
| C4 | prenotazione vaccino | Italy | 1/1/15 – 4/22/22 | Health | Web Search | URL-1 |
| C5 | tachipirina vaccino | Italy | 1/1/20 – 4/22/22 | Health | Web Search | URL-2 |
| C6 | brufen vaccino + ibuprofene vaccino | Italy | 1/1/20 – 4/22/22 | Health | Web Search | URL-3 |
| C7 | Nimesulide vaccino | Italy | 1/1/15 – 4/22/22 | Health | Web Search | URL-4 |

Such an investigation confirmed our suspicion (**Figure I3**). Indeed, the web search volumes are perfectly aligned with national vaccinations. Nimesulide-related RSV was negligible. This scenario is plausibly causal since substances such as paracetamol and ibuprofen are adopted to treat the mild and moderate adverse reactions of COVID-19 vaccines.

**Figure I3.**
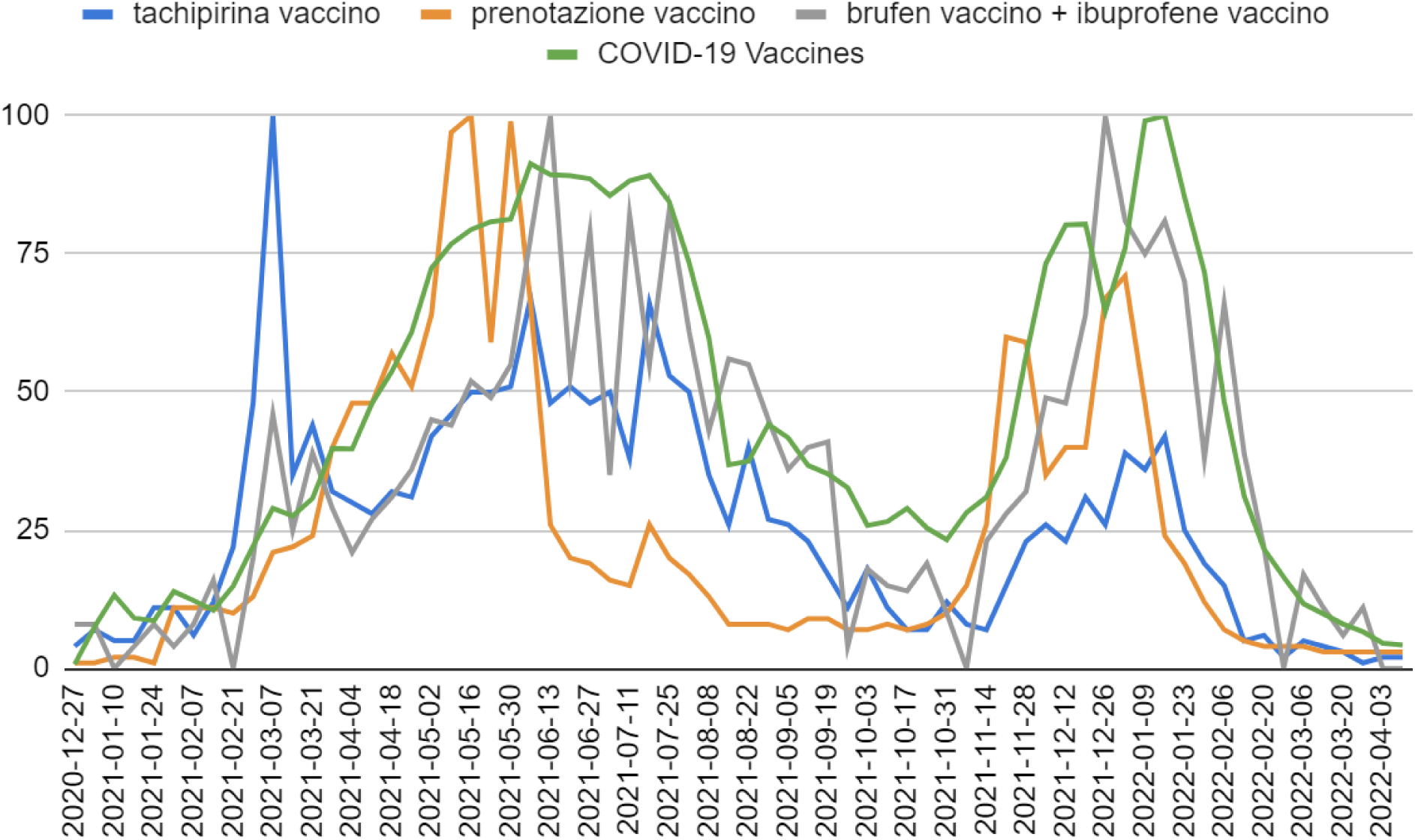
COVID-19 vaccinations and vaccine-related queries in Italy from January 2021 to April 2022. The volumes of each query have been downloaded separately and put in this graph (therefore, no quantitative information can be deduced from this graph). Nimesulide-related RSV was negligible.

The first peak of the query “tachipirina vaccino” (Tachipirina vaccine, March 2021) is probably due to the web interest in paracetamol as a possible substance to soothe the symptomatic manifestations deriving from vaccination (e.g., https://www.health.gov.au/sites/default/files/documents/2021/02/covid-19-vaccines---successivamente-alla-vaccinazione-contro-il-covid-19-after-your-covid-19-vaccination.docx). This indicates that media interest in drugs may cause RSV increases unrelated to epidemiological phenomena. Furthermore, this web interest seems to be linked to a potentially incorrect practice such as assuming paracetamol before vaccination (e.g., https://sip.it/2021/07/13/bisogna-assumere-paracetamolo-prima-di-sottoporsi-alla-vaccinazione-per-prevenire-gli-effetti-avversi/). Since the vaccination dataset was not normal (Shapiro-Wilk P=.004886), we calculated the Spearman correlation between COVID-19 vaccinations (x2), “tachipirina vaccino” query (y2), and “brufen vaccino + ibuprofene vaccino” query (z2) to confirm the above qualitative examinations, obtaining r_x2y2_=0.88, 95% CI: [0.78, 0.93], P_x2y2_<2.2e-16 and r_x2z2_=0.85, 95% CI: [0.72, 0.91], P_x2z2_=4.338e-16. The following vectors and commands were used (weekly values from 28 March 2021 to 10 April 2022, thus excluding the media clamor-related peak):

*x2 <-*

*c(39.8,39.7,47.9,53.7,60.8,72.4,76.8,79.4,80.8,81.3,91.3,89.3,89.2,88.6,85.6,88.2,89.2,84.4,73.4,59.8,36.8,37.5,44.2,41.7,36.7,35.2,32.7,25.9,26.6,29.0,25.4,23.3,28.2,31.0,38.2,56.5,73.3,80.2,80.4,64.6,76.1,99.1,100.0,85.3,71.7,48.3,31.3,21.6,16.5,11.7,9.9,8.0,6.6,4.6,4.2)*

*y2 <-*

*c(32,30,28,32,31,42,46,50,50,51,67,48,51,48,50,38,66,53,50,35,26,40,27,26,23,17,11,18,11,7,7,12, 8,7,15,23,26,23,31,26,39,36,42,25,19,15,5,6,2,5,4,3,1,2,2)*

*z2 <-*

*c(29,21,27,31,36,45,44,52,49,55,78,100,53,79,35,82,55,83,61,43,56,55,45,36,40,41,4,18,15,14,19,10,0,23,28,32,49,48,64,100,81,75,81,70,38,66,39,22,0,17,11,6,11,0,0)*

*shapiro.test(x2), cor.test(x2,y2,method=“spearman”,exact=FALSE), spearman.ci(x2, y2, nrep = 1000, conf.level = 0.95), cor.test(x2,z2,method=“spearman”,exact=FALSE)*, and *spearman.ci(x2, z2, nrep = 1000, conf.level = 0.95)*.

After that, we have downloaded the following keywords (**Table I4**):

**Table I4.**
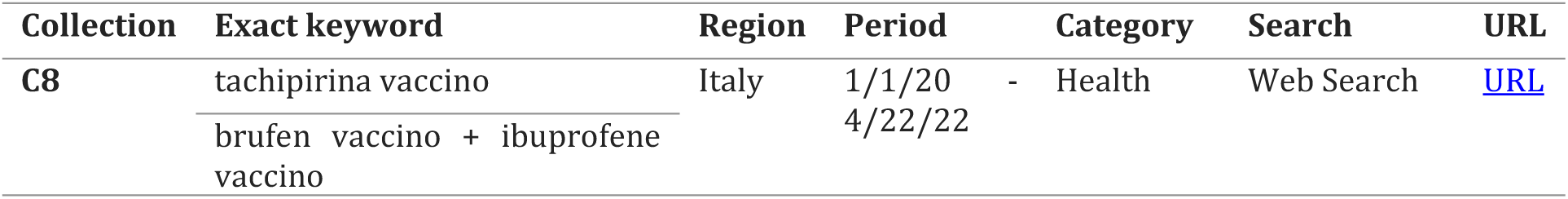
Paracetamol and ibuprofen vaccine-related exact queries.

By doing so, we were able to directly compare the search volumes of paracetamol and ibuprofen concerning COVID-19 vaccines from January 2020 to April 2022. Specifically, the Welch t-test returned P=6.193e-11. The average RSV were 16 (SD 19) for paracetamol (X2) and 3 (SD 4) for ibuprofen (Y2). The following vectors and commands were used:

*X2 <-*

*c(1,0,2,2,1,3,2,0.5,1,0.5,0,0.5,1,0.5,2,0.5,1,2,0,2,1,1,0.5,1,1,1,1,1,2,2,3,1,1,1,1,2,1,2,1,4,4,2,3,2,2,2,2,1,3,1,2,4,7,6,6,10,11,7,12,23,48,100,33,45,33,31,29,32,32,44,46,52,51,52,68,49,52,46,50,40,62,53,48,33,28,41,27,25,24,19,13,18,9,7,7,12,8,7,15,23,26,22,31,30,41,33,41,23,19,16,6,6,2,5,4,4,0,4,2,4)*

*Y2 <-*

*c(0,1,0,0,0,0,1,0,0,0,2,1,0,0.5,0,0,0,0,0,0.5,0,0,0,0,0,0,0,1,0,0,0,0,0,0,1,1,1,0,0,1,0,0,0,0,0,1,1,0,0,1,0,1,1,0,1,1,1,1,2,1,3,7,3,6,5,3,4,4,5,6,6,7,8,9,12,14,7,11,7,12,10,12,8,7,8,9,6,5,6,6,1,2,2,2,3,1,0,3,4,5,7,6,9,14,11,12,12,9,6,9,5,3,0,2,1,1,1,0,0,0)*

*t.test(X2, Y2, alternative = c(“greater”)), mean(X2), SD(X2), mean(Y2), SD(Y2), mean(Z2), SD(Z2)*.

Therefore, these findings suggest that paracetamol was the most used drug to counteract common COVID-19 vaccine-related side effects.

Regarding C3, the “Nimesulide” web topic registered a marked and significant increase from October 2020 onwards, preceded by a drop after the first COVID-19 wave (May – July 2020). Indeed, considering that the dataset showed no trend (**Figure I4**), the Welch t-test returned P=2.361e-07. The average values were 47 (SD 5) before October 2020 (x3) and 69 (SD 13) after October 2020 (y3). The following vectors and commands were used:

*x3 <-*

*c(49,43,44,43,46,38,41,55,41,54,45,40,44,45,43,41,41,41,42,46,39,47,49,54,46,44,41,46,46,45,48,47,45,56,50,51,56,48,45,51,42,46,45,47,46,57,48,46,52,50,50,47,43,46,43,46,47,50,50,54,54,51,61,47,37,42,41,50,46)*

*y3 <- c(53,62,82,69,59,74,100,58,64,59,73,54,67,60,81,95,61,68,78)*

*t.test(x3, y3, alternative = c(“less”)), mean(x3), sd(x3), mean(y3), sd(y3)*.

**Figure I4.**
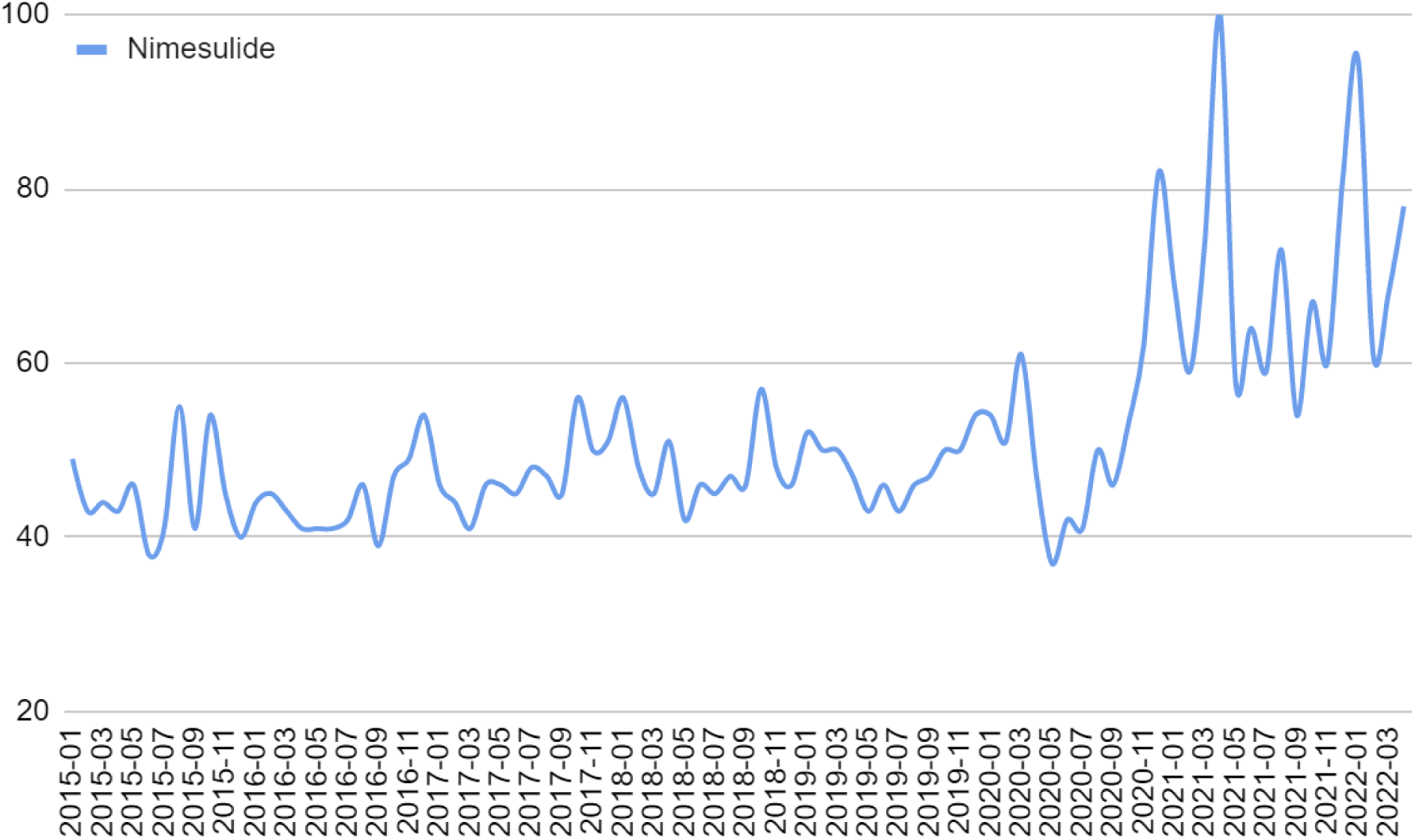
Nimesulide monthly web interest in Italy from January 2015 to April 2022.

Nonetheless, it is worth noting an unusual peak in April 2020 (first COVID-19 wave); indeed, the RSV reached its maximum value since 2015. Top and rising queries confirmed the direct association between these searches and COVID-19 (**Table I5**) during the pandemic period (**Table I6**).

**Table I5.**
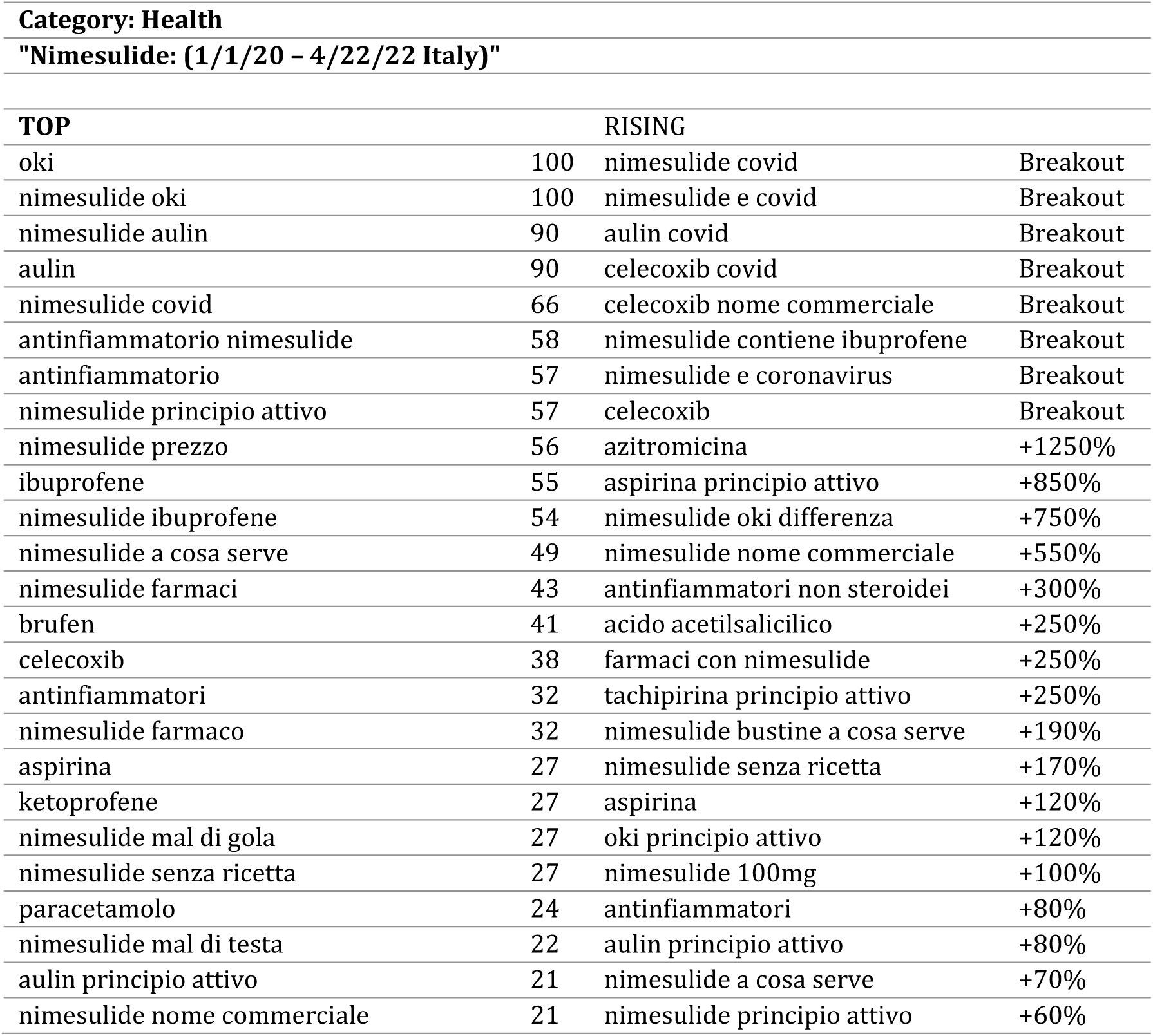
Nimesulide top and rising queries from January 2020 to April 2022.

**Table I6.** Nimesulide topic from January 2020 to April 2022.

| Collection | Exact keyword | Region | Period | Category | Search | URL |
| --- | --- | --- | --- | --- | --- | --- |
| C9 | Nimesulide (Drug) | Italy | 1/1/20 - 1/22/22 | Health | Web Search | URL |

Then, we check the “Fever” related queries before 2020 to ensure its causal relationship with seasonal flu (**Tables I7, I8**).

**Table I7.** Fever topic from January 2015 to December 2019.

| Collection | Topic | Region | Period | Category | Search | URL |
| --- | --- | --- | --- | --- | --- | --- |
| C10 | Fever (Medical condition) | Italy | 1/1/15 - 1/1/20 | Health | Web Search | URL |

**Table I8.** Fever top and rising queries from January 2015 to December 2019. * Possible confounding factors.

| Category: Health |  |  |  |
| --- | --- | --- | --- |
| Fever: (1/1/15 - 1/1/20, Italy) |  |  |  |
| TOP |  | RISING |  |
| febbre | 100 | influenza 2019 quanto dura la febbre | Breakout |
| la febbre | 28 | fluibron febbre e dolore | Breakout |
| febbre alta | 16 | febbre alta influenza 2019 | Breakout |
| febbre bambini | 15 | febbre dopo vaccino meningococco b* | Breakout |
| febbre sintomi | 10 | vaccino meningococco b febbre* | Breakout |
| tachipirina febbre | 6 | influenza 2017 quanto dura la febbre | Breakout |
| febbre alta bambini | 4 | influenza senza febbre 2016 | Breakout |
| febbre senza sintomi | 4 | influenza 2018 quanto dura la febbre | 3600% |
| rimedi febbre | 4 | sintomi febbre del nilo* | +800% |
| febbre nei bambini | 3 | nurofen febbre e dolore adulti | +350% |
| febbre 37 | 3 | febbre del nilo* | +300% |
| antibiotico febbre | 3 | nurofen febbre | +300% |
| temperatura febbre | 3 | come abbassare la febbre in modo naturale | +250% |
| abbassare la febbre | 3 | come capire se hai la febbre | +250% |
| febbre neonato | 2 | come abbassare la febbre | +200% |
| febbre e tosse | 2 | fever | +170% |
| farsi venire la febbre* | 2 | bronchite senza febbre | +170% |
| mal di gola e febbre | 2 | bentelan febbre | +140% |
| febbre dopo vaccino* | 2 | febbre 37.5 | +140% |
| fever | 2 | brividi senza febbre | +140% |
| febbre bassa | 2 | febbre e raffreddore | +130% |
| febbre 38 | 2 | febbre dopo antibiotico | +120% |
| misurare la febbre | 2 | febbre dopo vaccino | +110% |
| febbre persistente | 2 | febbre a 38 | +110% |
| febbre a 37 | 2 | nurofen bambini | +100% |

Although the association has been confirmed, we have detected potential confounding factors such as vaccines and other pathologies (e.g., West Nile Fever) or other practices (e.g., such as getting a fever voluntarily). For this reason, we looked for the trends of the aforementioned confounders (**Table I9**) and found that they did not influence the seasonal trends of the “seasonal flu – fever” relationship (**Figure I5**).

**Table I9.**
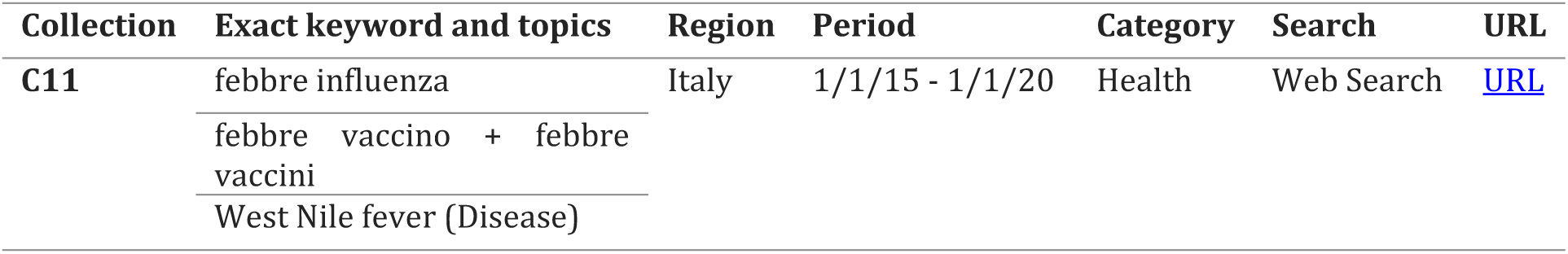
Fever-seasonal flu confounding factors from January 2015 to December 2019.

**Figure I5.**
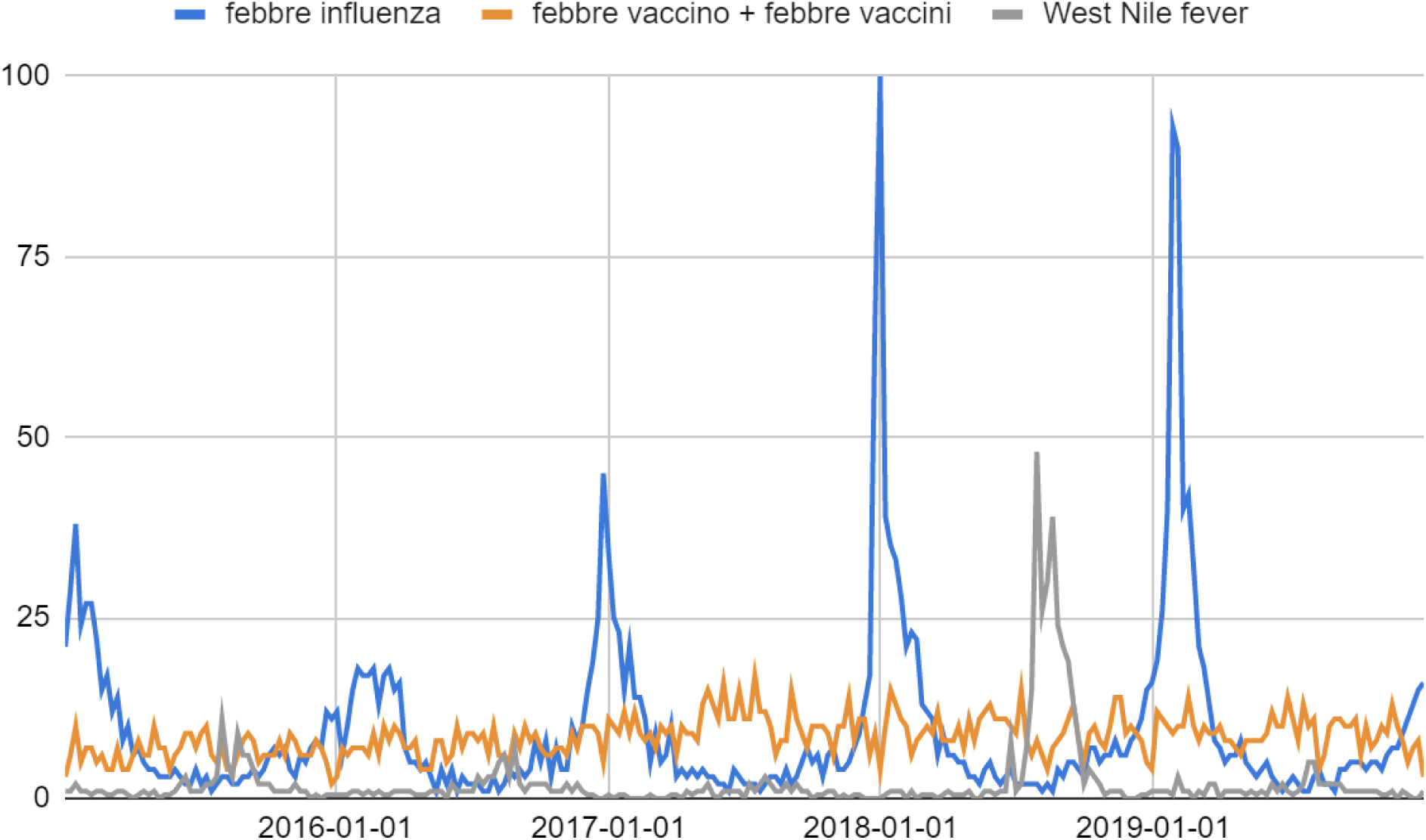
Fever-seasonal flu confounding factors RSV in Italy from January 2015 to December 2019.

## References

[1] Tennant JP, Ross-Hellauer T. The limitations to our understanding of peer review. Res Integr Peer Rev. 2020 Apr 30;5:6. doi: 10.1186/s41073-020-00092-1. PMID: 32368354; PMCID: PMC7191707.

[2] Fleming I. Who is afraid of being a reviewer? An A-Z of tips and tricks for peer review. Cardiovasc Res. 2021 Jul 7;117(8):e104–e105. doi: 10.1093/cvr/cvab180. PMID: 34198333.

[3] Bohannon J. Who’s afraid of peer review? Science. 2013 Oct 4;342(6154):60–5. doi: 10.1126/science.2013.342.6154.342_60. PMID: 24092725.

[4] Rinott E, Kozer E, Shapira Y, Bar-Haim A, Youngster I. Ibuprofen use and clinical outcomes in COVID-19 patients. Clin Microbiol Infect. 2020 Sep;26(9):1259.e5-1259.e7. doi: 10.1016/j.cmi.2020.06.003. Epub 2020 Jun 12. PMID: 32535147; PMCID: PMC7289730.

[5] Davis R, Brogden RN. Nimesulide. An update of its pharmacodynamic and pharmacokinetic properties, and therapeutic efficacy. Drugs. 1994 Sep;48(3):431–54. doi: 10.2165/00003495-199448030-00009. Erratum in: Drugs 1994 Nov;48(5):793. PMID: 7527762.

[6] Eccles R. Efficacy and safety of over-the-counter analgesics in the treatment of common cold and flu. J Clin Pharm Ther. 2006 Aug;31(4):309–19. doi: 10.1111/j.1365-2710.2006.00754.x. PMID: 16882099.

[7] Ministero della Salute, COVID-19. URL: https://www.salute.gov.it/portale/nuovocoronavirus/homeNuovoCoronavirus.jsp?lingua=english (Accessed 25 April 2022).

[8] Indolfi C, Spaccarotella C. The Outbreak of COVID-19 in Italy: Fighting the Pandemic. JACC Case Rep. 2020 Jul 15;2(9):1414–1418. doi: 10.1016/j.jaccas.2020.03.012. Epub 2020 Apr 1. Erratum in: JACC Case Rep. 2020 Aug;2(10):1656. PMID: 32835287; PMCID: PMC7270641.

[9] Rovetta A, Bhagavathula AS. The Impact of COVID-19 on Mortality in Italy: Retrospective Analysis of Epidemiological Trends. JMIR Public Health Surveill. 2022 Apr 7;8(4):e36022. doi: 10.2196/36022. PMID: 35238784; PMCID: PMC8993143.

[10] Eysenbach G. Infodemiology and infoveillance: framework for an emerging set of public health informatics methods to analyze search, communication and publication behavior on the Internet. J Med Internet Res. 2009 Mar 27;11(1):e11. doi: 10.2196/jmir.1157. PMID: 19329408; PMCID: PMC2762766.

[11] Giacchetta I, Primieri C, Cavalieri R, Domnich A, de Waure C. The burden of seasonal influenza in Italy: A systematic review of influenza-related complications, hospitalizations, and mortality. Influenza Other Respir Viruses. 2022 Mar;16(2):351–365. doi: 10.1111/irv.12925. Epub 2021 Oct 26. PMID: 34704361; PMCID: PMC8818820.

[12] Trentini F, Pariani E, Bella A, Diurno G, Crottogini L, Rizzo C, Merler S, Ajelli M. Characterizing the transmission patterns of seasonal influenza in Italy: lessons from the last decade. BMC Public Health. 2022 Jan 6;22(1):19. doi: 10.1186/s12889-021-12426-9. PMID: 34991544; PMCID: PMC8734132.

[13] Nuti SV, Wayda B, Ranasinghe I, Wang S, Dreyer RP, Chen SI, Murugiah K. The use of google trends in health care research: a systematic review. PLoS One. 2014 Oct 22;9(10):e109583. doi: 10.1371/journal.pone.0109583. PMID: 25337815; PMCID: PMC4215636.

[14] Stupple A, Singerman D, Celi LA. The reproducibility crisis in the age of digital medicine. NPJ Digit Med. 2019 Jan 29;2:2. doi: 10.1038/s41746-019-0079-z. Erratum in: NPJ Digit Med. 2019 Mar 19;2:19. PMID: 31304352; PMCID: PMC6550262.

[15] Greenland S. Invited Commentary: The Need for Cognitive Science in Methodology. Am J Epidemiol. 2017 Sep 15;186(6):639–645. doi: 10.1093/aje/kwx259. PMID: 28938712.

[16] Rovetta A. Reliability of Google Trends: Analysis of the Limits and Potential of Web Infoveillance During COVID-19 Pandemic and for Future Research. Front Res Metr Anal. 2021 May 25;6:670226. doi: 10.3389/frma.2021.670226. PMID: 34113751; PMCID: PMC8186442.

[17] Greenland S. Transparency and disclosure, neutrality and balance: shared values or just shared words? J Epidemiol Community Health. 2012 Nov;66(11):967–70. doi: 10.1136/jech-2011-200459. Epub 2012 Jan 19. PMID: 22268131.

[18] Cheung MW, Vijayakumar R. A Guide to Conducting a Meta-Analysis. Neuropsychol Rev. 2016 Jun;26(2):121–8. doi: 10.1007/s11065-016-9319-z. Epub 2016 May 21. PMID: 27209412.

[19] Saracco, A. Dr. Strangelove: Or How I Learned to Stop Worrying and Love the Citations. Math. Intell. 2022. doi: 10.1007/s00283-021-10146-x

[20] Our Health Service. Consigli post vaccino anti COVID-19 Comirnaty (Pfizer/ BioNTech). URL: https://www.hse.ie/eng/services/news/newsfeatures/covid19-updates/covid-19-vaccine-materials/covid-19-vaccine-translated-materials/pfizer-biontech-comirnaty-vaccine-aftercare-italian.pdf (Accessed 2 May 2022).

[21] Our Health Service. Dopo aver ricevuto il vaccino anti COVID-19 Moderna. URL: https://www.hse.ie/eng/services/news/newsfeatures/covid19-updates/covid-19-vaccine-materials/covid-19-vaccine-translated-materials/moderna-vaccine-aftercare-italian-.pdf (Accessed 2 May 2022).

